# Declining Opioid Distribution to US Hospitals

**DOI:** 10.1101/2021.05.03.21256543

**Authors:** Sarah A. Eidbo, Amalie K. Kropp Lopez, Joseph D. Hagedorn, Varkey Mathew, Daniel E. Kaufman, Kenneth L. McCall, Brian J. Piper

## Abstract

The United States is enduring a preventable opioid crisis, particularly involving a population specifically being treated in a hospital setting. This project analyzed trends in distribution of opioids by hospitals in the United States from 2000 to 2019. Opioids monitored included buprenorphine, codeine, fentanyl, hydrocodone, hydromorphone, meperidine, methadone, morphine, oxycodone, oxymorphone, powdered opium, remifentanil, and tapentadol. The Automation of Reports and Consolidated Orders System (ARCOS) reports controlled substances via the Drug Enforcement Administration (DEA). Data from ARCOS reports 5 and 7 from 2000 to 2019 were utilized for an observational study on hospital opioid distribution in the US. Morphine milligram equivalents (MME) were calculated using oral conversion factors for each opioid. The MME per person per state was calculated to compare data from the peak year, 2012, to data from 2019. Opioid use in the United States peaked in 2012 with a 46.6% decline from 2012 to 2019. Within the US, 25 of 50 states have seen a decrease of 50% or greater. Of the opioid compounds observed, buprenorphine has seen increased (123%) hospital use from 2000 to 2019. All other opioids have been experiencing a slow decline (>40%) in hospital use since that time. This study demonstrated the progress made thus far by hospitals in curbing the US opioid epidemic.

## INTRODUCTION

The United States (US) is currently enduring a persistent opioid crisis with 128 people dying each day from opioid overdoses [1]. The US Department of Health and Human Services estimates that 40% of opioid overdose deaths involve a prescription opioid [2]. Despite the US constituting only 4.4% of the global population, it constituted 30.2% of global opioid consumption [3]. Recent data from the American Hospital Association state that there are currently 6,146 US hospitals with 924,107 staffed beds [4]. In an observational study of opioid exposure and related adverse effects, opioids were utilized in treatment for over half of hospital admissions of nonsurgical patients in 286 US hospitals [5]. Opioids are closely linked with care within the hospital setting. Roughly 50 million surgical procedures a year are performed in the United States. Six percent of patients undergoing minor or major surgery continued opioid use 90 days after surgery, well after the normal scope of post-procedure pain treatment [6]. These findings align with the 5% incidence of new long-term opioid use after the first opioid exposure identified by the Oregon prescription drug monitoring program data, which includes non-operative opioid prescriptions [7]. The severe adverse effects that acute and chronic opioid use can cause are also important. Within the inpatient hospital setting, opioids are among the most frequently associated with potentially life-threatening events. Up to 14% of postoperative patients that were prescribed opioids experienced an adverse event. Patients were three times as likely to experience an adverse drug event as someone who was not prescribed opioids [8]. Among 300,000 surgeries, 12.2% of patients experienced an opioid-related adverse event, leading to greater overall costs, longer hospitalizations, and increased likelihood of readmission [9].

Despite the prevailing perception that the opioid epidemic is worsening, practitioners and hospitals have taken note of these trends [10]. Steps to hinder the epidemic are already taking effect. Recent hospital initiatives include stewardship programs for better education of prescribers as well as increased efforts to reduce negative outcomes – both aimed at decreasing the risk of opioid-related adverse events [11].

This observational study aimed to determine trends in hospital retail distribution of opioids in the US from 2000 to 2019, using the Automation of Reports and Consolidated Orders System (ARCOS). ARCOS is a comprehensive drug reporting system for substances controlled by the Drug Enforcement Administration (DEA), which reports on an array of opioids and opioid derivatives in addition to many other substances and has been examined in prior pharmacoepidemiological reports [12, 13, 14]. Several schedule II and III opioids used for pain control were studied using this database including codeine, oxycodone, hydromorphone, hydrocodone, meperidine, morphine, powdered opium, oxymorphone, remifentanil, tapentadol, and fentanyl. Two drugs utilized for also opioid use disorder (OUD), buprenorphine and methadone, were also analyzed.

## METHODS

### Procedures

All data was collected from ARCOS [15] for this observational study. The database is an ongoing reporting system that monitors DEA-controlled substances from their manufacture through their retail distribution to hospitals, pharmacies, practitioners, and teaching institutions, which are collectively grouped under business activities in several reports. Data from reports 5 and 7 were collected for 2000 to 2019. Report 5 details the average purchased amount of each drug, for each business activity, in each state. Report 7 gives the average purchased amount of drug for each business activity within the entire United States. The opioids included were buprenorphine, codeine, fentanyl base, hydrocodone, hydromorphone, meperidine, methadone, morphine, oxycodone, oxymorphone, powdered opium, and tapentadol. This study was deemed exempt by the IRBs of the University of New England and Geisinger.

### Statistics

The programs GraphPad Prism, Microsoft Excel, and JMP were used to graph and analyze this data. American Community Survey estimates of yearly state populations were used with opioids reported per 100 population [16]. To compare opioids of different formulations, established oral conversion factors were used to calculate the morphine milligram equivalent (MME) of each opioid listed with their corresponding multipliers. Oral conversion factors used were morphine: 1, oxycodone: 1.5, fentanyl: 75, hydrocodone: 1, hydromorphone: 4, oxymorphone: 3, tapentadol: 0.4, codeine: 0.15, meperidine: 0.1, methadone: 8, and buprenorphine: 10 [17]. The morphine milligram equivalent is the milligrams of morphine that the amount of opioid would be equivalent to. The percent change was calculated using the MME per person per state to compare 2012, the year with the highest US MME total, to years 2017-2019. State values that were outside a 95 percent confidence interval, calculated as mean +/-1.96 ^*^ Standard Deviation, were considered statistically significant.

## RESULTS

Due to the evolving nature of the data collected in this study, three separate percent changes were calculated as more recent data was made available by ARCOS. These three percent changes were between 2012 relative to 2017, 2018, and 2019. Hospital opioid use in the United States has seen a 31.1% decrease from 2012 to 2017 (Figure 1). When total opioid weights were calculated for the United States by year, the year 2012 had the highest hospital opioid distribution (19,188.91 kg MME). In 2017, the opioid kg MME was 13,227.88 kg MME, resulting in a 31.1% decrease. From 2012 to 2018, hospital opioid use saw a 39.7% decrease (Figure 1). The year 2012 was again used for the highest hospital opioid distribution by total opioid weight, with 19,188.91 kg MME. This was compared with the opioid kg MME of 2018, which was 11,574.61 kg MME. Between 2012 and 2019, hospital opioid use has seen a 46.6% change (Figure 1). When total opioid weights were calculated for the United States by year, 2012 was the year with the highest hospital opioid distribution, with 19,188.91 kg MME. When compared with the MME of 2019, 10,246.95 kg MME, a net decrease of 46.6% was seen.

**Figure 1.**
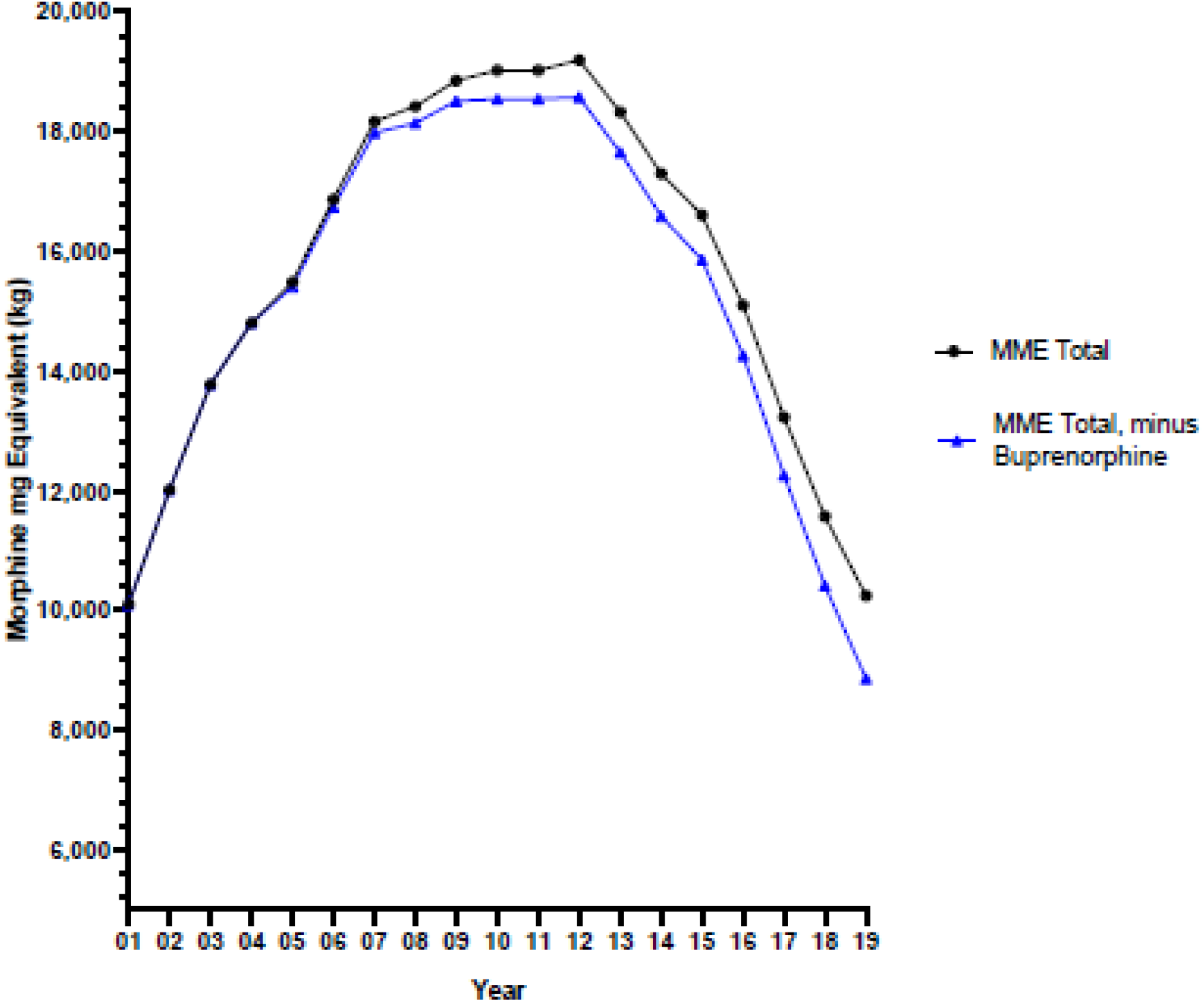
Total opioid distribution in morphine milligram equivalents (MME) in kilograms from 2001 to 2019 as reported by the Drug Enforcement Administration’s Automated Reports and Consolidated Orders System to hospitals for buprenorphine, codeine, fentanyl, hydrocodone, hydromorphone, meperidine, methadone, morphine, oxycodone, oxymorphone, and tapentadol (black) or with buprenorphine removed (blue).

When hospital opioid totals were broken down by their individual opioids, patterns became more apparent (Figure 2). A wide range in distribution weights amongst opioids required that they be separated into three groups. The opioids with the highest hospital distribution were oxycodone, morphine, and hydrocodone, all three of which displayed declining usage over the past several years. The middle group of opioids included codeine, methadone, hydromorphone, meperidine, buprenorphine and tapentadol. These drugs also displayed a steady decline, except for buprenorphine, which had distribution increases, and tapentadol, which rose after coming onto the market and then began to fall in 2017. The group of opioids with the lowest distribution included fentanyl, powdered opium, oxymorphone, and remifentanil. These drugs all remained steady.

**Figure 2.**
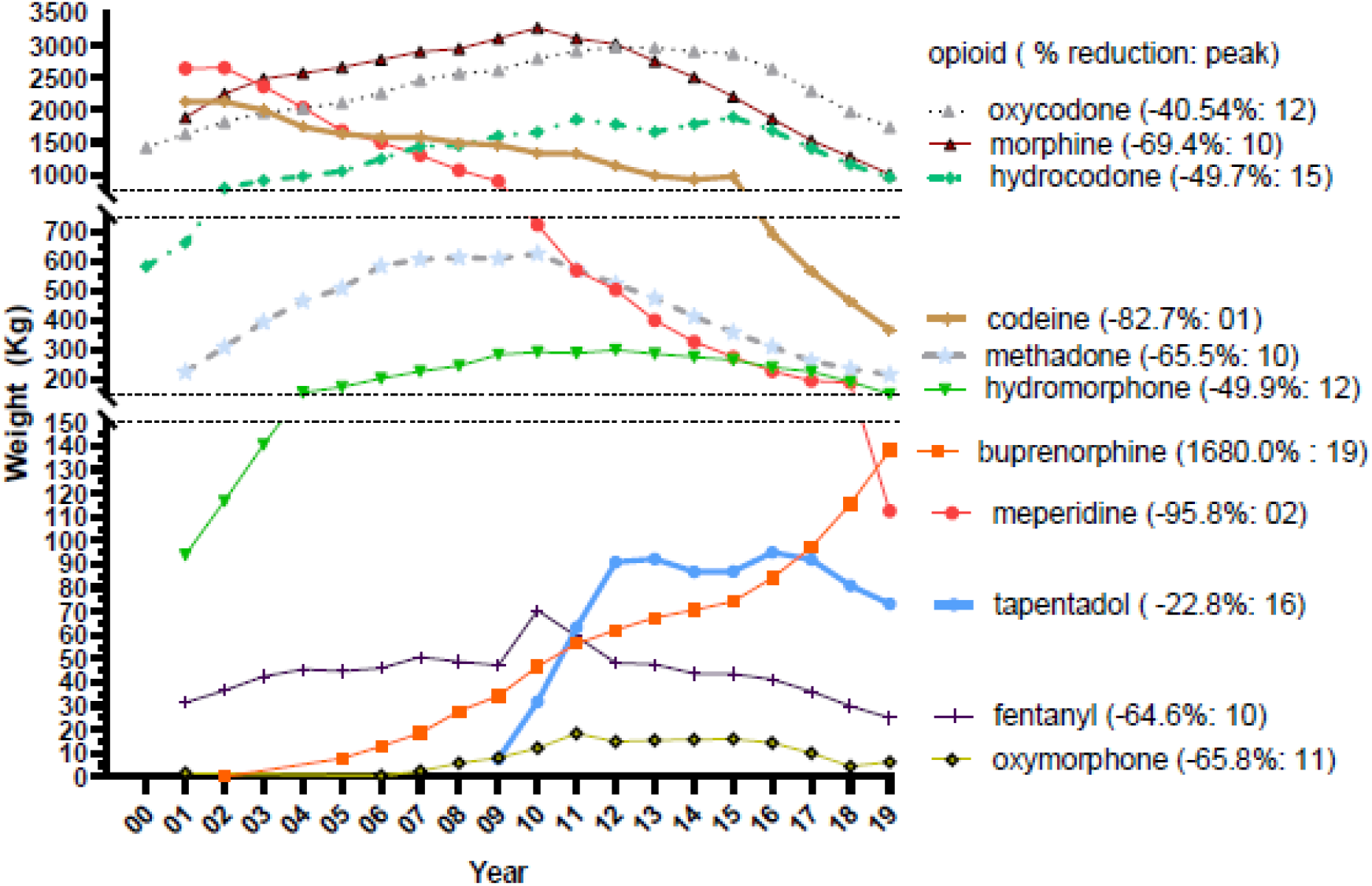
Individual opioid weights (kg) distributed to US hospitals as reported by the Drug Enforcement Administration’s Automated Reports and Consolidated Ordering System (ARCOS) from 2001 to 2018. The percent change relative to the peak year is shown in parentheses.

Opioids were then organized by their percent change between 2012, the peak year, and 2019 (Figure 3). All opioids displayed a percent decrease except for buprenorphine, which had a +123% increase. Six opioids had decreases of 50% or over: hydromorphone (−49.9%), oxymorphone (−57.7%), methadone (−58.7%), morphine (−66.9%), codeine (−67.5%), and meperidine (−77.6%).

**Figure 3.**
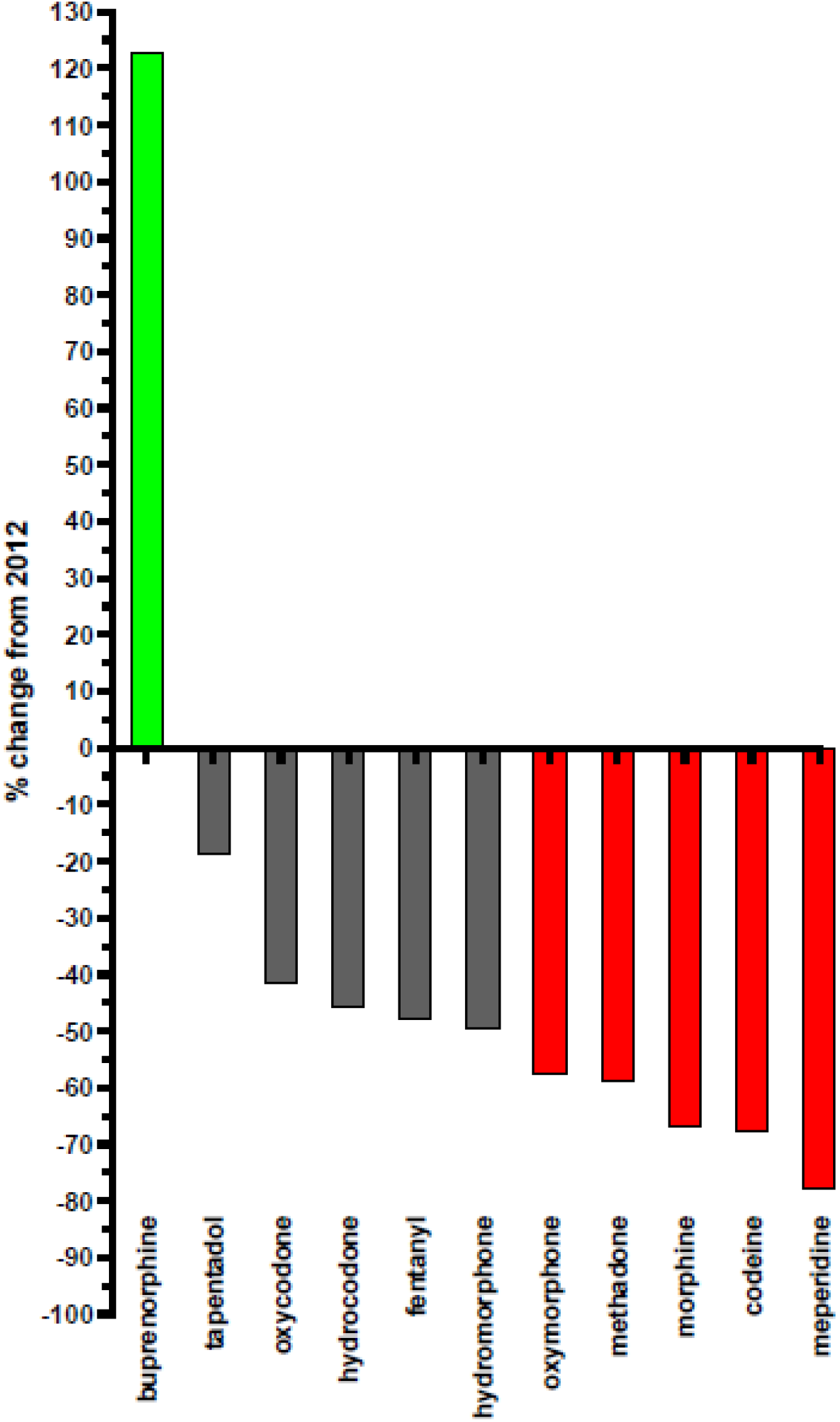
Individual opioid percent change as reported by the Drug Enforcement Administration’s Automated Reports and Consolidated Ordering System (ARCOS) between 2012 and 2019. Changes of greater than +50% are shown in green and greater than -50% in red.

Hospital opioid distribution was then broken down by state, by the opioid weight distributed per person by each state’s population (Figure 4, 5). The average opioid weight distributed per person was 17.91 (SD= 6.92) mg. Compared with 2012, several changes are worth noting – firstly, that half of the United States experienced a 50% or greater percent decrease in hospital-distributed opioid weights per person (Figure 7). Secondly, the state with the highest hospital-distributed opioid weight per person in 2012 was South Carolina, which has seen a 94% decrease in 2019. The percent decrease that South Carolina experienced was calculated to be an outlier among the states’ percent decreases. It is also worth mentioning that in 2019, the states with the highest hospital opioid distribution per person by weight were Alaska, Montana, Colorado, and North Dakota (Figure 5). These have changed from 2012, when the states with the highest distribution were South Carolina, Alaska, North Dakota, and South Dakota (Figure 6). Alaska and North Dakota have remained among the states with the highest hospital opioid distributions by weight per person (Figure 5).

**Figure 4.**
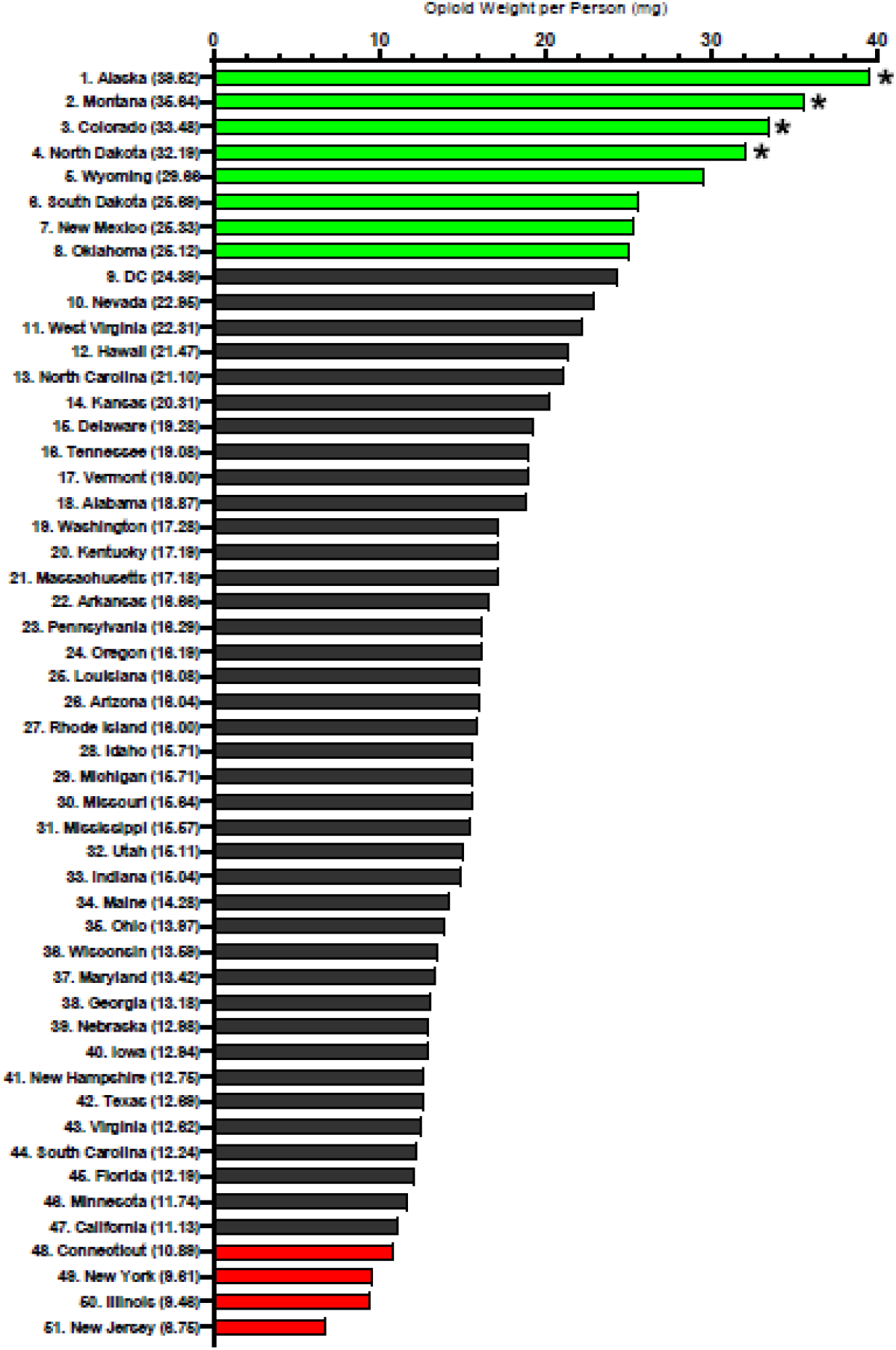
Hospital-distributed opioid weights per person per state as reported by the Drug Enforcement Administration’s Automated Reports and Consolidated Ordering System (ARCOS) in milligrams in 2019. Average opioid weight per person was 17.91mg, standard deviation was 6.92mg. States with opioid weights per person of over 24.84mg were one standard deviation above the average weight and are colored green. States with opioid weights per person of under 10.98mg were one standard deviation below the average weight and are colored red. States with an opioid weight per person that were calculated to be outliers are marked with asterisks (p < .05) and include Alaska, Montana, Colorado, and North Dakota.

**Figure 5.**
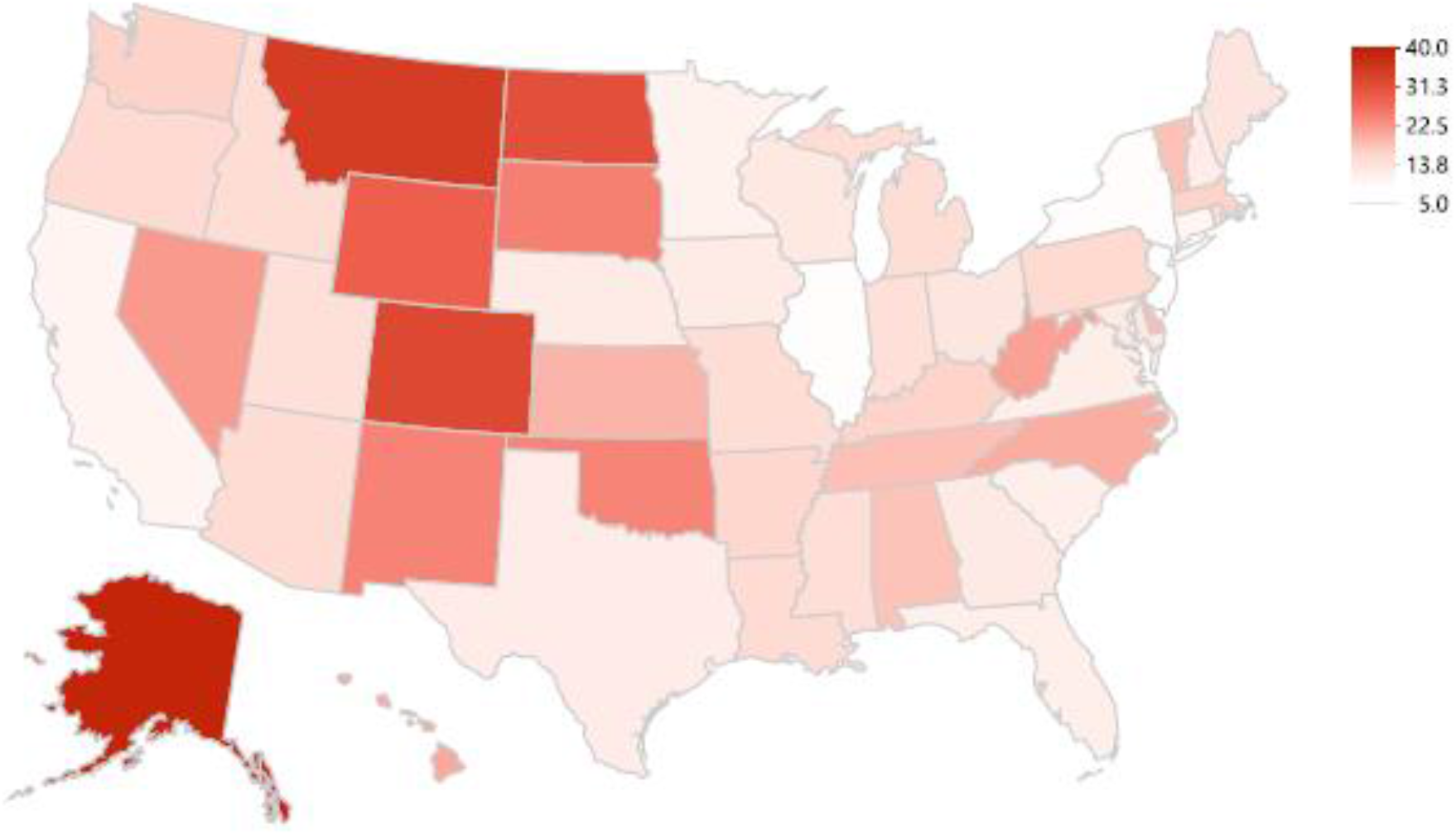
Hospital-distributed opioid weights as reported by the Drug Enforcement Administration’s Automated Reports and Consolidated Ordering System (ARCOS) in milligrams per person per state in 2019.

**Figure 6.**
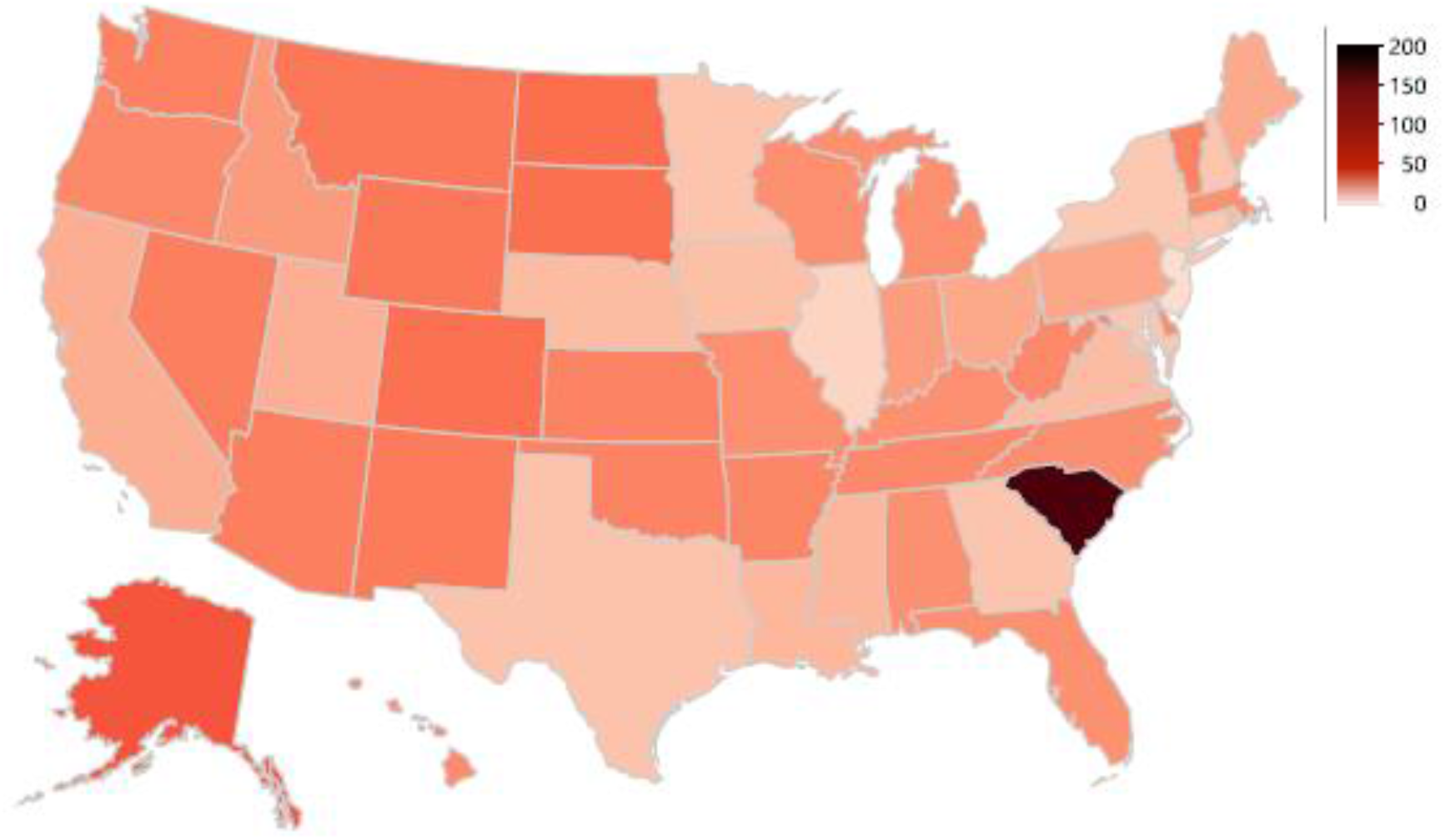
Hospital-distributed opioid weights as reported by the Drug Enforcement Administration’s Automated Reports and Consolidated Ordering System (ARCOS) in milligrams per person per state in 2012.

**Figure 7.**
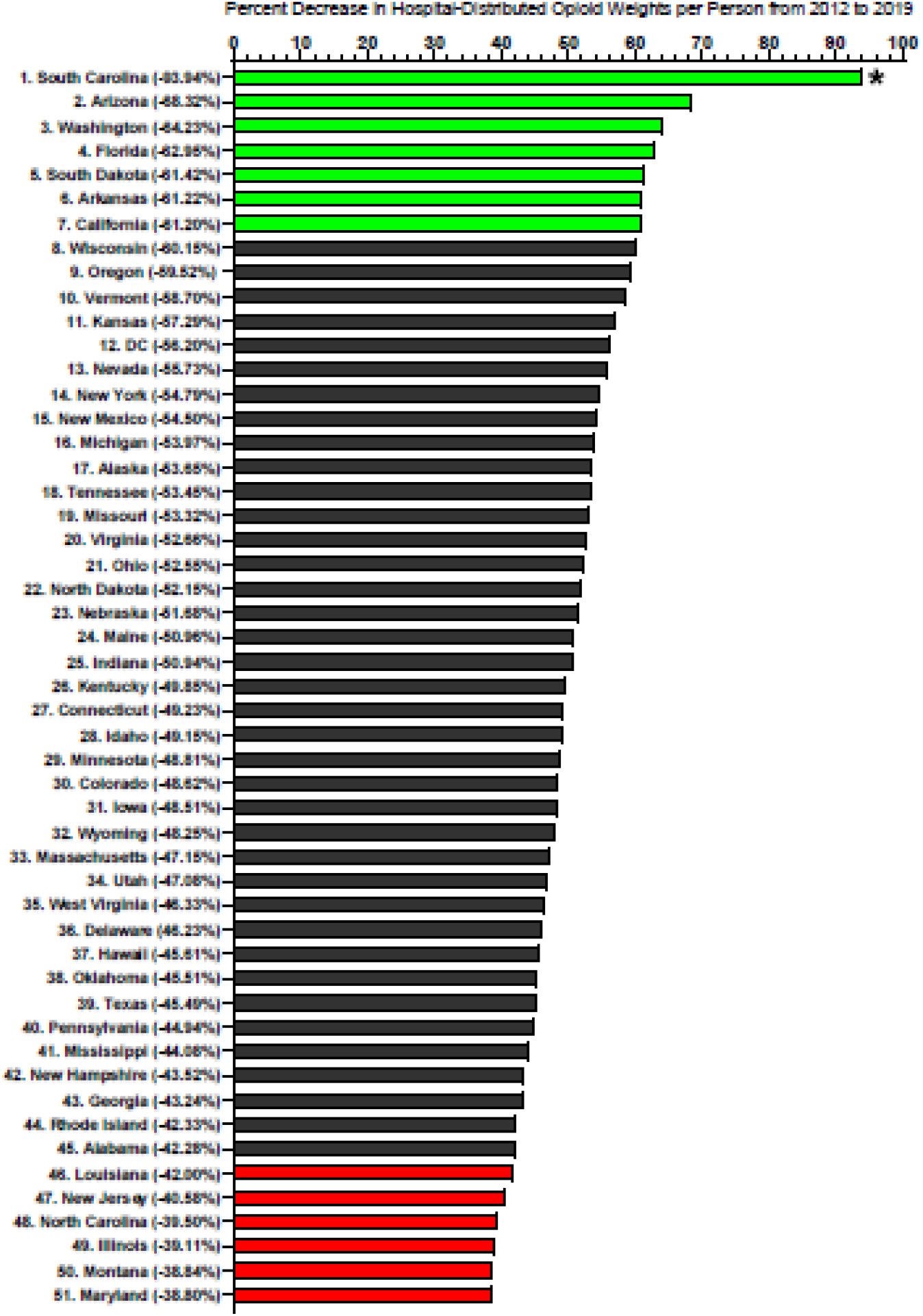
Percent decrease in hospital-distributed opioid weights as reported by the Drug Enforcement Administration’s Automated Reports and Consolidated Ordering System (ARCOS) per person per state in milligrams from 2012 to 2019. Average decrease in opioid weight per person was -51.38%, and the standard deviation was 9.34%. States with decreases in opioid distributions per person of more than - 69.68% were one standard deviation beyond the average decrease and shown in green. States with decreases in opioid distributions per person of under -33.08% were one standard deviation below the average decrease and are shown in red. States with a decrease in opioid distribution per person that were calculated to be outliers are marked with asterisks (p < .05).

## DISCUSSION

This study identified the changes in opioid distribution within United States hospitals from 2000 to 2019. Many opioid medications have been declining in use since peaking in 2012. Hospital opioid distribution overall has seen a decrease of 46.6% between 2012 and 2019. This comes with the exception of buprenorphine, which has seen a 123% increase in use since 2012, as it is being used to combat opioid use disorder. Other opioid medications observed in this investigation reached their peak distributions in 2011 with declining rates since.

The disproportionate amounts of certain opioid medications distributed in some states is another interesting find. In 2012, South Carolina had the largest hospital-distributed opioid weight per person in the country – it was over twice the weight per person of the state with the second highest distributed opioid weight. Further investigation revealed this high weight to be mostly due to codeine and hydrocodone distribution. Other opioid medications appeared to be within a similar range of the distributed weights of other states. Although the reasoning behind the distribution of these two opioid drugs in South Carolina is unclear, it had receded by 2019. South Carolina had a remarkable 94% decrease in hospital-opioid distribution per person, making it 44^th^ in rank for the highest opioid distribution weight.

In 2019, Alaska had the highest hospital distribution per person followed by Montana, Colorado, and North Dakota. Within Alaska, the rates of opioid-related inpatient hospitalizations were 28.5 per 100,000 persons in 2016 and 26.0 per 100,000 persons in 2017, with total inpatient hospitalization charges exceeding $23 million [18]. The state is beginning to implement changes to reduce the escalating overdose deaths and high hospitalization rates.

The lowest hospital opioid distribution per person in 2019 was seen in New Jersey, followed by Illinois, New York, and Connecticut. In several of these states, guidelines have been implemented that include a multifaceted approach to reducing hospital opioid distribution. These approaches include performing a patient evaluation before prescribing, getting informed consent for opioid treatment, and periodic review of the complete medical record. This comes from state advisory guidelines on use of controlled substances in treatment of intractable pain [19]. The opioid epidemic affects people of all metrics across the nation, but these state-dependent differences indicate that further investigation is necessary for better understanding of this complex crisis.

The US is not alone in the observation of this trend over the last two decades. In Canada, one out of five adults reports living with chronic pain. Codeine, oxycodone, and hydromorphone were also the most commonly prescribed opioid medications for pain in Canada. However in a study from 2013 to 2016 in Ontario, Saskatchewan and British Columbia, some slight decreases in opioid prescriptions were observed. The investigation revealed that fewer Canadians were being prescribed opioids (8.0% fewer) and fewer citizens are starting opioid therapy as treatment for pain in all age groups (9.6% fewer). Changes were also made to prescribed opioid dosages and durations. In summary, frequency in prescribing opioids for under a week increased and frequency of opioids prescribed for longer than a week declined [20].

Hospitals and insurance companies across the US have been searching for methods to decrease the number of opioid medications prescribed to patients. This multifaceted problem must be addressed according to multiple variables. Funding has been allocated to several areas thus far: addiction prevention strategies, programs aimed at identifying individuals at high risk of opioid use and misuse, increasing treatment facility number, and increased efforts of law enforcement agencies [21]. However, despite new safety nets put in place, physicians still balance on a fine line between treating the pain of a patient and potentially over-prescribing these complicated medications.

Hospitals in the United States are integrating improved guidelines to reduce amounts of potentially unnecessary opioid prescriptions. A recent report from 81 hospitals found that 98% implemented changes to their opioid delivery practices between July of 2017 and July of 2018. The changes made included improved prescriber education and new technologies to monitor prescribing practices, which limited the dosage and quantities of given opioids [22].

Post-surgical opioid use could be a potential gateway to chronic opioid use, if opioid medications are used incorrectly. With the low number of patients taking their prescribed doses of opioid medications after surgery, this suggests that perhaps their pain could be managed via other modalities. Patients’ pain can often be controlled with nonsteroidal anti-inflammatory drugs (NSAIDs) and acetaminophen, removing the need for opioids and other stronger pain medications. However, the decision to prescribe a patient NSAIDs and acetaminophen in the place of opioids and other strong pain medications comes down to the healthcare team. The healthcare team’s ability to educate patients and create appropriate expectations for home pain management is vital in these situations [23].

Certain strengths and weaknesses of this novel dataset should be taken into consideration. ARCOS only reports in the hospital business activity on agents that are sent by distributors to hospitals. Prescriptions that are written within the hospital – such as those following a surgery – but filled in a non-hospital pharmacy are reported by ARCOS as pharmacy-distributed opioids. However, the comprehensive nature of ARCOS makes it an important source for follow-up reports.

In conclusion, this study observed the differences and trends of 13 different opioids within hospitals from 2000 to 2019 at a national and state level. These changes support those observed at a national level for all opioid distribution - not just within hospitals. Since 2011, there has been a steady decrease in the distribution of opioids for pain and increases in buprenorphine for opioid use disorder. Hospitals were prescribing less opioids, while also taking steps to make prescribing opioids safer for patients. This is without a doubt a positive step toward combating the effects of the opioid crisis in the US healthcare system.

## Data Availability

Raw data is publicly available

https://www.deadiversion.usdoj.gov/arcos/retail_drug_summary/index.html

## Acknowledgements

Stephanie Nichols, PharmD provided feedback on an earlier version of this report.

